# Optimizing stress in breaking bad news: a randomized controlled trial on the psychophysiological effects of stress arousal reappraisal and worked-example interventions among medical students

**DOI:** 10.1101/2025.01.21.25320844

**Authors:** Michel Bosshard, Sissel Guttormsen, Urs Markus Nater, Felix Schmitz, Patrick Gomez, Christoph Berendonk

## Abstract

Breaking bad news (BBN) is among the most distressing communication tasks in the medical field, wherein physicians disclose serious diagnoses to their patients. Under stress, physicians may resort to maladaptive communication behaviors, potentially affecting patient’s health in the long-term. Therefore, it is essential to support medical professionals in effectively managing their stress responses early in their careers. Using the biopsychosocial model of challenge and threat as theoretical framework, we employed a 2 x 2 study design to examine the effects of stress arousal reappraisal (SAR; i.e., reinterpretation of bodily changes as functional coping resources) and worked example (WE; i.e., step-by-step demonstration of how to BBN) interventions on demand and resource appraisals and cardiovascular responses of 229 medical students engaged in simulated BBN encounters. Participants who prepared with WE reported more coping resources relative to demands after the BBN encounter than participants not preparing with WE. Participants receiving SAR instructions exhibited improved cardiovascular responses during the BBN task, indicated by increased cardiac output and decreased total peripheral resistance, than participants not receiving SAR instructions. These findings align with the notion that both interventions facilitate a shift from a threat to a challenge state, supporting their potential for integration into BBN training.

## Introduction

Stress is a ubiquitous phenomenon in the medical workplace and education^1^. Physicians are expected to manage high workloads under time pressure, while navigating challenges of illness, mortality, and the emotional upheaval experienced by their patients. *Breaking bad news* (BBN) represents an indispensable task, wherein physicians disclose serious diagnoses to their patients, often involving life-altering circumstances (e.g., stillbirth, cancer diagnosis^2,3^). The emotional responses of patients to bad news are unpredictable, ranging from silence and withdrawal to expressions of anger and disbelief^2,4^. At the same time, physicians often experience anxiety regarding BBN, with such encounters evoking acute stress responses^5–9^. Importantly, increased sympathetic arousal has been observed already among medical students in simulated BBN encounters (e.g.,^10,11^; see^8^ for a review). Physiological stress responses may divert the medical students’ attention away from the immediate clinical task towards stress management^12^. Consequently, elevated stress levels can impair their performance in educational and clinical settings (e.g., communication^13^). In BBN consultations, stress may result in suboptimal communication behaviors, such as providing false hope by either withholding the true severity of the diagnosis or through excessive optimism regarding the treatment^12,14,15^. While existing stress management programs in medical education have shown promising outcomes^16^, significant time commitment and stigmatization present obstacles to their implementation into medical training. Thus, there is a need for low-threshold approaches that facilitate efficient coping of students with stress at an early stage. To this end, the current study investigated the effects of *stress arousal reappraisal* (SAR) and *worked example* (WE)-based learning interventions on the psychophysiological stress response of medical students tasked with BBN. The *biopsychosocial model* (BPSM) *of challenge and threat* served as guiding theoretical framework.

### The biopsychosocial model of challenge and threat

The BPSM of challenge and threat applies to motivated performance situations, which require an active response of the individual and are driven by self-relevance, such as job interview, academic examinations, and public speaking^16–19^. While an active response is central in these contexts, it is the degree of self-relevance that determines the degree of task engagement^20^. According to theory (e.g.,^21,22^), task engagement is a prerequisite for challenge and threat states and is indicated by increased heart rate (HR) and decreased pre-ejection period (PEP, period between the start of left ventricular contraction and the opening of the aortic valve) when compared to rest periods. In the context of motivated performance situations, a psychological evaluation of situational demands and personal coping resources precedes the emergence of distinct cardiovascular response patterns^23,24^. Both psychological and cardiovascular stress responses can be conceptualized along a bipolar spectrum of challenge vs. threat^17,22^, allowing for meaningful interpretation of relative differences in responses.

A challenge-oriented stress response arises when perceived personal resources (e.g., knowledge, skills, familiarity) are sufficient to cope with the situational demands (e.g., difficulty, uncertainty, danger^17,22^). In contrast, a threat-oriented response emerges when the situational demands are perceived to exceed available resources. The resources-demands differential (also referred to as “Demand Resource Evaluation Score” ^25^) thus represents a psychological assessment that indicates challenge and threat on a continuous scale^20^. This evaluation is an automatic and dynamic process, meaning it can change as the situation progresses^26,27^. Importantly, the evaluation of personal resources is amenable to interventions (e.g.,^28–30^).

Regarding the cardiovascular system, an increase in sympathetic arousal caused by task engagement is presumed in both challenge and threat states. However, in a challenge state, the increase in sympathetic arousal is typically paired with higher cardiac output (CO; liters of blood pumped by the heart per minute) and lower total peripheral resistance (TPR; an index of vasoconstriction vs. vasodilation) than in a threat state, resulting in improved blood flow and oxygen distribution^31,32^. CO and TPR can be combined to create a continuous cardiovascular index of challenge and threat (standardized CO − standardized TPR^33^). A relatively higher cardiovascular index reflects relatively greater challenge or lesser threat. In some studies, stroke volume (SV; volume of blood pumped by the ventricle per beat) has been utilized as indicator of challenge and threat in place of CO, with higher SV representing challenge (e.g.,^19,34–36^). Cardiovascular indices of challenge have been linked to more positive health outcomes, while indices of threat have been associated with more negative health outcomes (e.g.,^37,38^).

### Stress arousal reappraisal

Stress is often viewed as inherently negative (i.e., distress), with the experience of arousal, such as an increased HR, seen as a hindrance to task performance in stressful situations. Consequently, many stress management techniques primarily focus on downregulating, reducing, or ignoring stress reactions altogether^39^. In contrast, SAR interventions emphasize the advantageous aspects of stress by educating individuals on the functionality of physiological arousal, framing it as beneficial for successful task performance (e.g.,^40–43^). For instance, rather than perceiving a fast and strong heartbeat as debilitating, individuals are encouraged to view it as a source of additional oxygen supply, enhancing performance in demanding situations. Thus, SAR interventions encourage individuals to embrace the body’s stress arousal, carrying the potential for individuals to primarily focus on the immediate task. In alignment with the BPSM of challenge and threat, by viewing stress arousal as a resource, a shift from threat-oriented to challenge-oriented responses can occur. Research indicates that, compared to control conditions, SAR leads to an increased resource evaluation and a more favorable resources-demands differential, while demand evaluation alone remains largely unaffected (e.g.,^28, 44^). In terms of cardiovascular measures, SAR leads to higher CO, lower TPR (^29,45,46^; but see^47^), and a higher cardiovascular index of challenge and threat (^43,44^; but see^21,47^) compared to control conditions.

### Worked example-based learning

WE-based learning is grounded in cognitive load theory, which posits that during learning, limited cognitive resources are allocated to cope with different types of cognitive load (i.e., intrinsic, extrinsic, germane^48^). Successful skill acquisition can be fostered by minimizing extrinsic load—inefficient use of cognitive resources due to poorly structured and presented information, so that working memory capacity is freed up for intrinsic (effective) load, arising from the difficulty of the information itself. In accordance with cognitive load theory, WEs provide step-by-step demonstrations of how to perform a complex task, thereby reducing extrinsic cognitive load imposed by the design of instructional materials^49^. WEs are most effective for novice learners^50^, as they enable schema acquisition^51^. Schemas facilitate the organization of related elements and can be retrieved as single units from memory. This is particularly valuable in high-stress situations, where cognitive resources may be sparse^52^. Within the BPSM of challenge and threat, knowledge and skills are integral to resource evaluation^17^. In social situations, well-learned tasks promote challenge-oriented cardiovascular responses compared to unlearned tasks (e.g.,^53^). Therefore, we propose that WE interventions can promote challenge states by supporting the successful acquisition of BBN skills, although WEs have not been previously examined within the BPSM of challenge and threat^54^.

### Current study

The goal of the current study was to evaluate the effectiveness of SAR and WE-based learning interventions on the psychophysiological stress response of medical students tasked with BBN to simulated patients (i.e., actresses trained to portray a real patient). Based on the outlined theory, we hypothesized that both SAR and WE interventions would improve the psychophysiological stress response, promoting a more challenge-type response. Our primary endpoints to differentiate between challenge and threat states were changes in the resources-demands differential and the cardiovascular index of challenge and threat, measured from baseline to post-BBN task and BBN task, respectively. We addressed the following main hypotheses:

1. Effects of the SAR intervention

1.1) Students receiving the SAR intervention exhibit a significantly more positive/less negative change in their resources-demands differential from baseline to post-BBN task than students not receiving the SAR intervention.
1.2) Students receiving the SAR intervention exhibit a significantly more positive/less negative change in their cardiovascular index of challenge and threat from baseline to BBN task than students not receiving the SAR intervention.
2. Effects of the WE-based learning intervention

2.1) Students receiving the WE intervention exhibit a significantly more positive/less negative change in their resources-demands differential from baseline to post-BBN task than students not receiving the WE intervention.
2.2) Students receiving the WE intervention exhibit a significantly more positive/less negative change in their cardiovascular index of challenge and threat from baseline to BBN task than students not receiving the WE intervention.

Additionally, HR and PEP reactivity from baseline to the BBN task were used to assess whether participants were actively engaged in the task. We expected a significant increase in HR and a significant decrease in PEP for the entire sample.

Furthermore, in line with the analytical approach taken in similar studies (e.g.,^35,44,45^), we analyzed secondary outcomes to gain a more nuanced understanding of the psychophysiological effects. We expected that the SAR and WE groups would show significantly more adaptive changes in resource evaluation (higher), CO (higher), TPR (lower), and SV (higher) compared to their respective control groups. No significant differences in the demand evaluation were expected across groups.

## Methods

This study has been preregistered on ClinicalTrials.gov (NCT05037318, 08/09/2021) and was approved by the cantonal ethics committee in Bern (2021-02098). Informed consent was obtained from all participants prior to the experimental procedure. The study was conducted in accordance with the World Medical Association’s Declaration of Helsinki, the ICH Good Clinical Practice (GCP) guidelines, and the Swiss Federal Human Research Act. We adhered to the CONSORT reporting guidelines^55^. A study protocol has been published^56^. In this paper, we focus on the cardiovascular and psychological indices of challenge and threat, which were assessed alongside other parameters (see^56^ for details).

### Sample size calculation

The sample size was calculated a priori with G*Power 3 software^57^. Based on the review of published data on the psychophysiological effects of the interventions, we concluded that an effect size of *d* = 0.4^58^ was reasonable and practically relevant. Given an alpha of .05 (two-tailed), *n* = 50 participants per group (*N* = 200) were necessary to test our hypotheses with a targeted statistical power of 0.80.

### Participants

A total of *N* = 229 third-year medical students participated in the study. To represent a broad range of Swiss students, recruitment emails were sent to four universities across different regions of Switzerland. M.B. oversaw participant recruitment, which began in April 2022 and continued until the target sample size was achieved in February 2024. Over half of the participants were from the University of Bern (*n* = 127), with the remaining students coming from the University of Basel (*n* = 44), the University of Fribourg (*n* = 39) and the University of Zurich (*n* = 19). The mean age of the participants was *M* = 22.42 years (*SD* = 1.83) with a mean BMI of *M* = 22.32 (*SD* = 2.79). Sixty-nine percent of the students were female (*n* = 158), of whom half used hormonal contraceptives (*n* = 79). Twenty-six percent of the participants were doing shift work (*n* = 59). All participants completed the entire experimental procedure and received remuneration of 150 Swiss Francs, plus travel expenses. To be eligible for study participation, medical students had to be enrolled in the third year at a Swiss university and be fluent in German. Third-year students have already completed basic communication courses but do not yet have specialized knowledge in BBN and thus represent an ideal population to test our hypotheses. We excluded students who reported suffering from cardiovascular or neuroendocrine conditions, using medication or psychoactive drugs known to affect the outcomes of interest, or wearing a pacemaker. Further, female participants could not be pregnant or lactating and—when possible—were tested in the first week after their menstruation to control for the potential effects of the fluctuating sexual hormones on the psychophysiological stress responses^59,60^.

### Study design

To test our hypotheses, we employed a 2 (SAR vs. No-SAR) × 2 (WE vs. No-WE) between-subjects design. Participants were stratified by sex and randomized with equal allocation ratio (within blocks of sizes 4 and 8) to one of the following conditions: (1) SAR-only, (2) WE-only, (3) SAR & WE or (4) No-intervention. M.B. generated a randomized allocation sequence in R, which was then concealed in an excel file, with the date and time of access of each allocation being logged. The participants were unaware of the four different conditions, whereas the simulated patients interacting with them did not know about the assigned condition. The experimenters were responsible for setting up the learning modules (i.e., interventions) and consequently were not blinded. The study was conducted in German.

### Study procedure

Medical students interested in the study completed an entry questionnaire, which was used to assess their eligibility and collect sociodemographic data.

Eligible students were invited to participate in an individual experimental session at the Institute for Medical Education in Bern, scheduled from 2pm to 4pm to control for circadian rhythm. They were instructed to restrain from alcohol consumption and intense physical activity 24 hr before the session, from heavy meals and caffeine consumption for 2 hr, and tobacco and food consumption for 1 hr prior to participation. Two participants reported that they did not comply with these requirements. The statistical analysis was rerun without these participants, which did not significantly impact any of the findings.

Upon arrival, the experimenter explained to the participants that they would first undergo baseline recordings, then learn how to BBN and apply their skills in a simulated BBN scenario, and finally, answer a series of questionnaires. Participants were informed that their BBN encounter would be recorded and rated with regard to the quality of their performance. After the experimental procedure was explained, written consent was obtained.

Sensors for the cardiovascular recording were attached and baseline cardiovascular activity was recorded for 5 min. Participants then read an introductory sentence to the BBN task, describing the prenatal setting. Afterwards, they were asked to evaluate their demands and resources for a first time. At this point, participants also evaluated their perceived skills in BBN, and reported on previous experiences and interest in BBN, and their motivation to perform well in the BBN task. This was followed by a web-based learning module of 40 min (see Fig. 1), during which participants received an introduction to BBN, and engaged in the BBN SPIKES protocol (Setting, Perception, Invitation, Knowledge, Emotions, Strategy & Summary^2^) and their respective interventions (SAR-only, WE-only, SAR & WE, or No-intervention) at their own pace. The order of the learning elements was identical for all participants: 1) introduction to BBN, 2) SPIKES protocol, 3) WE (for participants in the WE groups), 4) SAR or respective control screencast. The whole module was accessible at once, and participants were instructed to complete the module in the order given. Once they had completed the module, they could use the remaining time to navigate freely through the individual elements. After the learning period, participants evaluated again their demands and resources (post-intervention), received specific instructions about the diagnosis and had 5 more min to prepare for the communication task (see “BBN Task”). The simulated patient then knocked on the door, entered the room, and took a seat opposite the participant.

**Figure 1.**
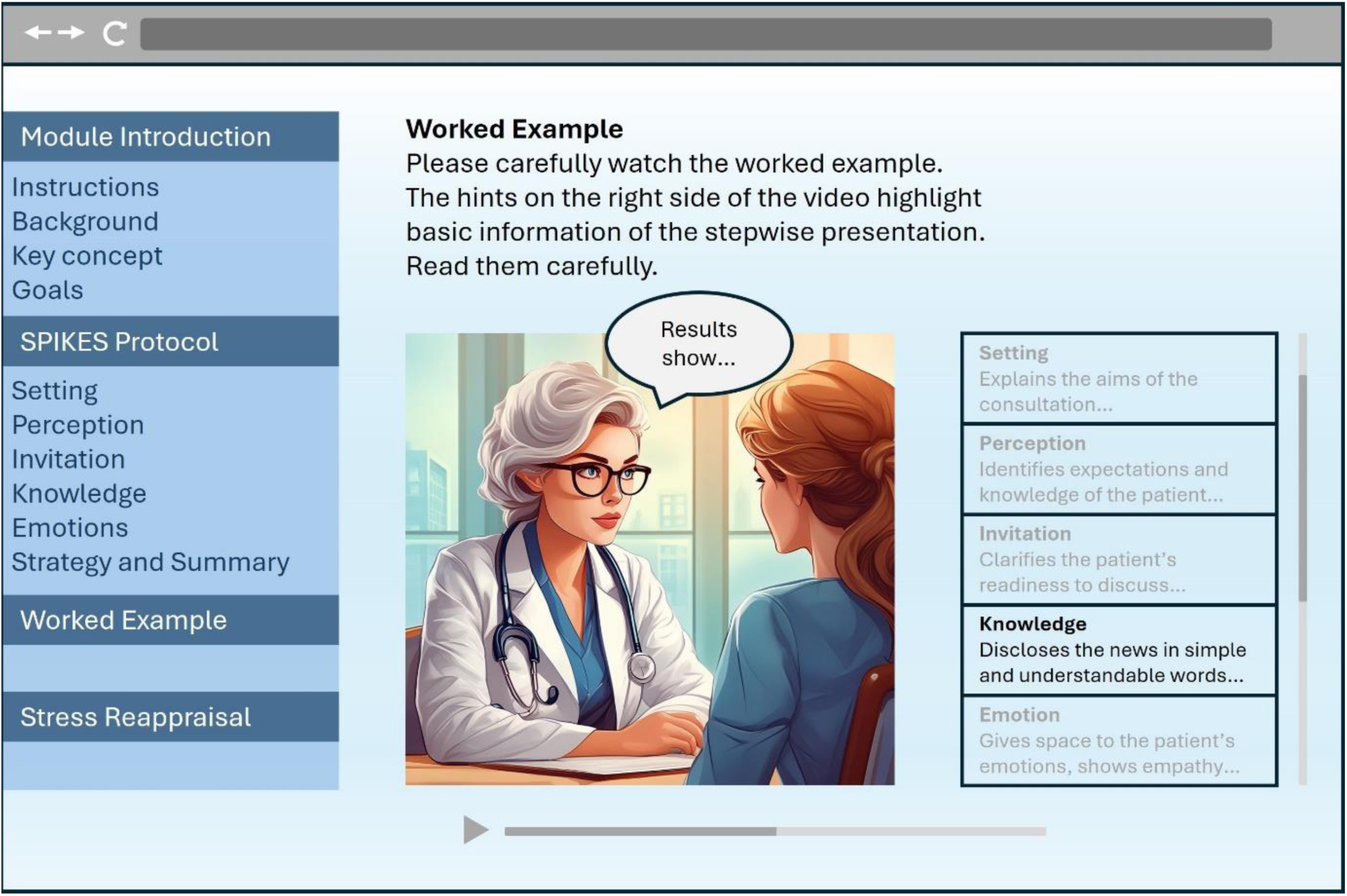
Illustration of the learning module. The worked example and stress arousal reappraisal interventions were only available for the respective groups.

Participants were informed that they had a maximum of 12 min for the BBN consultation. On average, the BBN consultation took 8 min and 28 s (*SD* = 1 min and 54 s). After the encounter, the simulated patient left the room. The cardiovascular activity was recorded 2 min prior, during, and 2 min after the BBN encounter. Following the task, participants retrospectively evaluated their experienced demands and resources (post-BBN) and answered the DASS-21 questionnaire. After the sensors were removed, participants were debriefed on the exact aims of the study, and any remaining questions were answered. Throughout the experiment, participants were seated at a table. The experimenter was present 1) at the beginning to explain the procedure and fit the cardiovascular devices, 2) before the learning module to pause cardiovascular recording and start the module, 3) after the learning module to resume cardiovascular recording and start video recording, 4) following the BBN encounter to stop video recording, and 5) at the end to remove the sensors and conduct the debriefing.

### BBN task

In the chosen scenario, the student took the role of a junior doctor, and the simulated patient portrayed a pregnant woman. The students’ task was to disclose a Trisomy 21 diagnosis appropriately, complying with the SPIKES protocol^2^. BBN in prenatal care is an important and growing challenge in clinical practice, that can affect the prospects of a family^61^. Due to the large number of participants, 11 simulated patients portrayed the role. The simulated patients received identical and detailed training from a professional instructor to ensure comparable performance. The role of the simulated patient was to display a state of shock and an inability to process further information after receiving the diagnosis.

### Measurements

#### Sociodemographic data

The variables age, sex, shift work, and affiliated university were obtained from the entry questionnaire, and the body mass index (BMI) was calculated for each participant at the beginning of the experimental session. These variables were treated as potential control variables^62–64^. In addition, the eligibility criteria regarding cardiovascular and neuroendocrine diseases, pacemaker use, medication, psychoactive drugs, and year of enrollment were assessed. Female participants also indicated the timing of their menstrual cycle, and whether they were pregnant, lactating, or using hormonal contraceptives. The latter was also treated as a potential control variable^65^.

#### Cardiovascular measures

The Finometer (FMS Finapres Medical Systems, Amsterdam, The Netherlands) recorded beat-to-beat blood pressure. For this purpose, a finger-cuff was wrapped around the middle phalanx of the left middle finger. Hydrostatic height correction of the finger relative to the heart level was active during measurement. The Finometer generates waveform measurements that resemble intra-arterial recordings and reconstructs brachial arterial pressure^66–68^. The Finometer has proven to measure changes in blood pressure accurately and precisely (e.g.,^69,70^).

The VU-Ambulatory Monitoring System^71,72^ (VU-AMS, Free University, Amsterdam, The Netherlands) was used to measure HR, PEP, and SV. This involved placing five electrodes on the thorax and two electrodes on the back for the recording of the electrocardiogram (ECG) and thorax impedance^73^.

Our primary endpoint, the cardiovascular index of challenge and threat, was computed by subtracting standardized TPR from standardized CO so that higher values represent more challenge-type cardiovascular responses. CO was determined by multiplying SV by HR, and TPR was calculated by dividing mean arterial pressure by CO.

#### Perceived demands and resources

Demand and resource evaluations were assessed before the BBN task with the items “How demanding do you expect the BBN task to be?” and “How able are you to cope with the demands of the BBN task?”, respectively. Participants answered two similar items after the task (“How demanding was the BBN task?” and “How able were you to cope with the demands of the BBN task?”). The items were measured on a 6-point Likert scale, ranging from 1 *not at all* to 6 *extremely*. We adapted the items from previous research^25,74,75^. We subtracted the demand score from the resource score to obtain the primary outcome resources-demands differential, which can range from -5 to +5, with higher scores indicating more challenge-type responses.

#### Depression, anxiety, and stress

Depression, anxiety, and stress were gauged on the DASS-21^76,77^, with 7 items per affective state (e.g., depression “I couldn’t seem to experience any positive feeling at all”; anxiety “I was worried about situations in which I might panic and make a fool of myself”; stress “I found myself getting upset rather easily”). Participants answered the items by referring to the past week and using a scale ranging from 0 *Did not apply to me at all* to 3 *Applied to me very much or most of the time*. Total scores of each subscale can range from 0 to 21, with higher values representing a stronger experience of the affective state. In this study, the scales possessed reasonable to good internal consistency (McDonald’s ω: depression 0.81, anxiety 0.74, stress 0.83). Depression, anxiety, and stress were used to characterize the sample and evaluated as potential control variables^78^.

#### BBN experience, BBN skills, BBN interest, and BBN motivation

Participants stated on a *yes*/*no* item if they had experience in BBN and rated their perceived BBN skills (1 *very low* to 7 *very high*), their general interest in the topic of BBN (1 *very little interest* to 7 *very interested*), and motivation in performing well on the BBN task (1 *not at all motivated to perform well* to 7 *absolutely motivated to perform well*). These items were used to characterize the sample and treated as potential control variables.

### Interventions

The learning module consisted of the well-established SPIKES protocol^2^, supplemented by the interventions according to group assignment (see Fig. 1).

#### Stress arousal reappraisal

The content of the intervention was based on SAR research^21,28–30,35,44^ and presented as a 7-min screencast. The screencast stated that experiencing stress arousal is normal and shows that a person cares about the task at hand. Further, the functionality of various bodily stress responses (e.g., an increased HR facilitating oxygen supply) was illustrated within an evolutionary framework. As part of the intervention, participants were instructed to reframe stress as performance enhancing rather than debilitating and to reflect on past and future stressful experiences. The corresponding control material consisted of a 7-min screencast discussing the neurocognitive processes involved in memory (similar to^21^). The SAR intervention and the control material are available in the OSF repository (https://osf.io/9aqwn/).

#### Worked Example

The WE consisted of a 10-min video showing a physician engaging in BBN with a simulated patient. In this video, the physician followed the SPIKES protocol^2^, providing applied examples for each step with accompanying textual hints (see^79^). For example, the physician initiated the step *Knowledge* by prefacing the disclosure of the diagnosis with an announcement of bad news (“Unfortunately, I do not have any good news for you today…”). Participants assigned to the corresponding control condition instead were given 10 additional min to repeat the SPIKES protocol.

### Reduction of the cardiovascular data

We computed the mean over the entire duration of each of the four cardiovascular measurement periods (5-min baseline, 2-min pre-task, BBN task, 2-min post-task). The Beatscope software was used to analyze the data from the Finometer. Due to device malfunctioning, 3 recordings were missing completely, for 12 participants the periods of interest were only partially recorded (e.g., Finometer stopped after first minute of BBN), and for 2 participants the recording quality was insufficient. We used the VU Data Analysis & Management Software to analyze the VU-AMS recordings. First, divergent interbeat intervals were examined in the ECG, and abnormal R-peaks were either corrected or labelled as artefacts. Second, ensemble averaged impedance-and electro-cardiograph complexes for the four periods were scored. Twelve VU-AMS recordings were missing completely and in 11 cases, we were unable to reliably score the recordings due to noisy data (e.g., detached electrodes). In total, complete cardiovascular data of 189 participants remained for the statistical analysis.

### Statistical analysis

The statistical analysis was conducted using R^80^ (version 4.3.1), with R-packages *nlme*^81^ (version 3.1.164) and *emmeans*^82^ (version 1.10.0). For all tests, a significance alpha of .05 was used. Group differences in sociodemographic and BBN-related variables were assessed with univariate analysis of variance (for continuous variables) or chi-squared tests (for categorical variables). Variables with significant group differences were included in the statistical models of the main outcomes as control variables. As shown in Supplementary Table S1, we found only a significant group effect for BBN motivation.

For all outcomes, we calculated linear mixed effects regressions based on restricted maximum likelihood estimation, with degrees of freedom based on the containment method of the *nlme* package^83^. To evaluate the changes from baseline, two models were tested for each outcome. Model 1 included fixed effects for SAR (SAR/No-SAR), WE (WE/No-WE), time, and for the control variable BBN motivation (centered around the grand mean), as well as the two-way interactions SAR × time, WE × time, SAR × WE, and BBN motivation × time. Model 2 was based on Model 1 and additionally tested the three-way interaction SAR × WE × time. For the task engagement parameters HR and PEP, an additional Model 0 without any interactions was further computed to test the main effect of time. For the cardiovascular outcomes, time had 4 levels (baseline, pre-BBN, BBN, post-BBN), whereas for the demand and resource outcomes, it had 3 levels (baseline, post-intervention, post-BBN). In our analysis, we focused on the changes from baseline to subsequent periods (e.g., from baseline to BBN) and did not examine the overall omnibus effect of time. Hypotheses were specifically tested on the change from baseline to BBN for the cardiovascular index of challenge and threat, and from baseline to post-BBN for the resources-demands differential. To interpret significant three-way interactions (e.g., SAR × WE × BBN), we conducted four post-hoc tests examining two-way interactions at each level of the other intervention (i.e., SAR x BBN at WE = 0 and WE = 1; WE x BBN at SAR = 0 and SAR = 1).

Models with varying random effect structures were compared based on AIC (Akaike Information Criterion) and BIC (Bayesian Information Criterion). All models included a random intercept for participants to accommodate for individual variances. Nesting participants in universities and/or simulated patients did not improve the models. A heterogeneous residual variance structure was preferable for all outcomes except for the cardiovascular index of challenge and threat.

Model assumptions regarding linearity, residual homoscedasticity, and normality were adequately met based on visual evaluation of residuals vs. fitted value plots, QQ-plots of the residuals, and random effect plots.

## Results

### Sociodemographic and BBN-related variables

Results for the sociodemographic and BBN-related variables are reported in Supplementary Table S1. Only the motivation to perform well in the BBN task differed significantly among the experimental groups. The SAR-only group was significantly more motivated than the WE-only group (*p* = .005). There were no significant differences in motivation between the other groups. The same results were found for the sub-sample with complete cardiovascular data (*N* = 189).

### Primary outcomes

Descriptive statistics for the primary outcomes are reported in the Supplementary Tables S2 and S3, whereas the estimated linear mixed effects regression models are reported in Tables 1 and 2.

**Table 1.**

| Fixed Effect | Model 1 |  |  | Model 2 |  |  |
| --- | --- | --- | --- | --- | --- | --- |
|  | Beta | SE | *p* | Beta | SE | *p* |
| Intercept | -0.86 | 0.16 | < .001 | -0.85 | .17 | < .001 |
| SAR <sup>a</sup> | -0.01 | 0.22 | .98 | -0.01 | 0.25 | .97 |
| WE <sup>b</sup> | -0.08 | 0.22 | .72 | -0.09 | 0.25 | .71 |
| Post-Intervention <sup>c</sup> | -0.22 | 0.15 | .14 | -0.22 | 0.17 | .20 |
| Post-BBN <sup>c</sup> | -0.69 | 0.18 | < .001 | -0.73 | 0.21 | < .001 |
| Motivation <sup>d</sup> | 0.09 | 0.11 | .42 | 0.09 | 0.11 | .42 |
| SAR × WE | -0.19 | 0.27 | .48 | -0.16 | 0.35 | .65 |
| SAR × Post-Intervention | 0.18 | 0.18 | .33 | 0.17 | 0.25 | .50 |
| SAR × Post-BBN | 0.37 | 0.22 | .087 | 0.46 | 0.31 | .14 |
| WE × Post-Intervention | 0.18 | 0.18 | .31 | 0.17 | 0.25 | .49 |
| WE × Post-BBN | **0.62** | **0.22** | **.005** | 0.70 | 0.30 | .022 |
| Motivation × Post-Intervention | 0.15 | 0.12 | .20 | 0.15 | 0.12 | .20 |
| Motivation × Post-BBN | 0.08 | 0.14 | .58 | 0.08 | 0.14 | .58 |
| SAR × WE × Pre-BBN |  |  |  | 0.01 | 0.35 | .97 |
| SAR × WE × Post-BBN |  |  |  | -0.17 | 0.43 | .70 |

Linear mixed effects regressions for the resources-demands differential. Significant
interaction effects ( $p < .05$ ) are presented in bold. <sup>a</sup> No-SAR = 0, SAR = 1. <sup>b</sup> No-WE = 0, WE
= 1. <sup>c</sup> Reference = baseline. <sup>d</sup> Motivation is grand mean centered.

**Table 2.**

| Fixed Effect | Model 1 |  |  | Model 2 |  |  |
| --- | --- | --- | --- | --- | --- | --- |
|  | Beta | *SE* | *p* | Beta | *SE* | *p* |
| Intercept | -0.11 | 0.27 | .69 | 0.01 | 0.27 | .98 |
| SAR<sup>a</sup> | 0.08 | 0.38 | .84 | -0.16 | 0.39 | .69 |
| WE<sup>b</sup> | 0.10 | 0.38 | .80 | -0.13 | 0.39 | .73 |
| Pre-BBN<sup>c</sup> | -0.15 | 0.16 | .33 | -0.28 | 0.18 | .11 |
| BBN<sup>c</sup> | -0.43 | 0.16 | .007 | -0.63 | 0.18 | < .001 |
| Post-BBN<sup>c</sup> | -0.25 | 0.16 | .11 | -0.37 | 0.18 | .038 |
| Motivation<sup>d</sup> | -0.07 | 0.18 | .71 | -0.07 | 0.18 | .71 |
| SAR × WE | 0.10 | 0.51 | .84 | 0.58 | 0.55 | .30 |
| SAR × Pre-BBN | 0.20 | 0.18 | .27 | 0.47 | 0.26 | .068 |
| SAR × BBN | **0.90** | **0.18** | **< .001** | 1.31 | 0.26 | < .001 |
| SAR × Post-BBN | 0.35 | 0.18 | .062 | 0.60 | 0.26 | .020 |
| WE × Pre-BBN | 0.11 | 0.18 | .56 | 0.37 | 0.25 | .15 |
| WE × BBN | -0.02 | 0.18 | .89 | 0.38 | 0.25 | .13 |
| WE × Post-BBN | 0.09 | 0.18 | .61 | 0.34 | 0.26 | .18 |
| Motivation × Pre-BBN | -0.02 | 0.12 | .86 | -0.02 | 0.12 | .86 |
| Motivation × BBN | 0.06 | 0.12 | .60 | 0.06 | 0.12 | .60 |
| Motivation × Post-BBN | 0.00 | 0.12 | .98 | 0.00 | 0.12 | .97 |
| SAR × WE × Pre-BBN |  |  |  | -0.54 | 0.36 | .14 |
| SAR × WE × BBN |  |  |  | **-0.84** | **0.36** | **.020** |
| SAR × WE × Post-BBN |  |  |  | -0.51 | 0.36 | .16 |

Linear mixed effects regressions for the cardiovascular index of challenge and threat.
Significant interaction effects ( $p < .05$ ) are presented in bold. <sup>a</sup> No-SAR = 0, SAR = 1. <sup>b</sup> No-
WE = 0, WE = 1. <sup>c</sup> Reference = baseline. <sup>d</sup> Motivation is grand mean centered.

#### Resources-demands differential

In line with hypothesis 2.1, Model 1 revealed a significant WE × Post-BBN effect, indicating that the change from baseline to post-BBN was significantly more positive for the WE groups (*M* = 0.11, *SE* = 0.15) than the No-WE groups (*M* = -0.50, *SE* = 0.15). Regarding hypothesis 1.1, the SAR × Post-BBN interaction was trending, indicating that the difference in changes from baseline to post-BBN between the SAR groups (*M* = -0.01, *SE* = 0.16) and No-SAR groups (*M* = -0.38, *SE* = 0.15) was in the expected direction. The Model 1 estimated resources-demands differentials across time are illustrated in Fig. 2. Model 2 revealed no significant three-way interactions.

**Figure 2.**
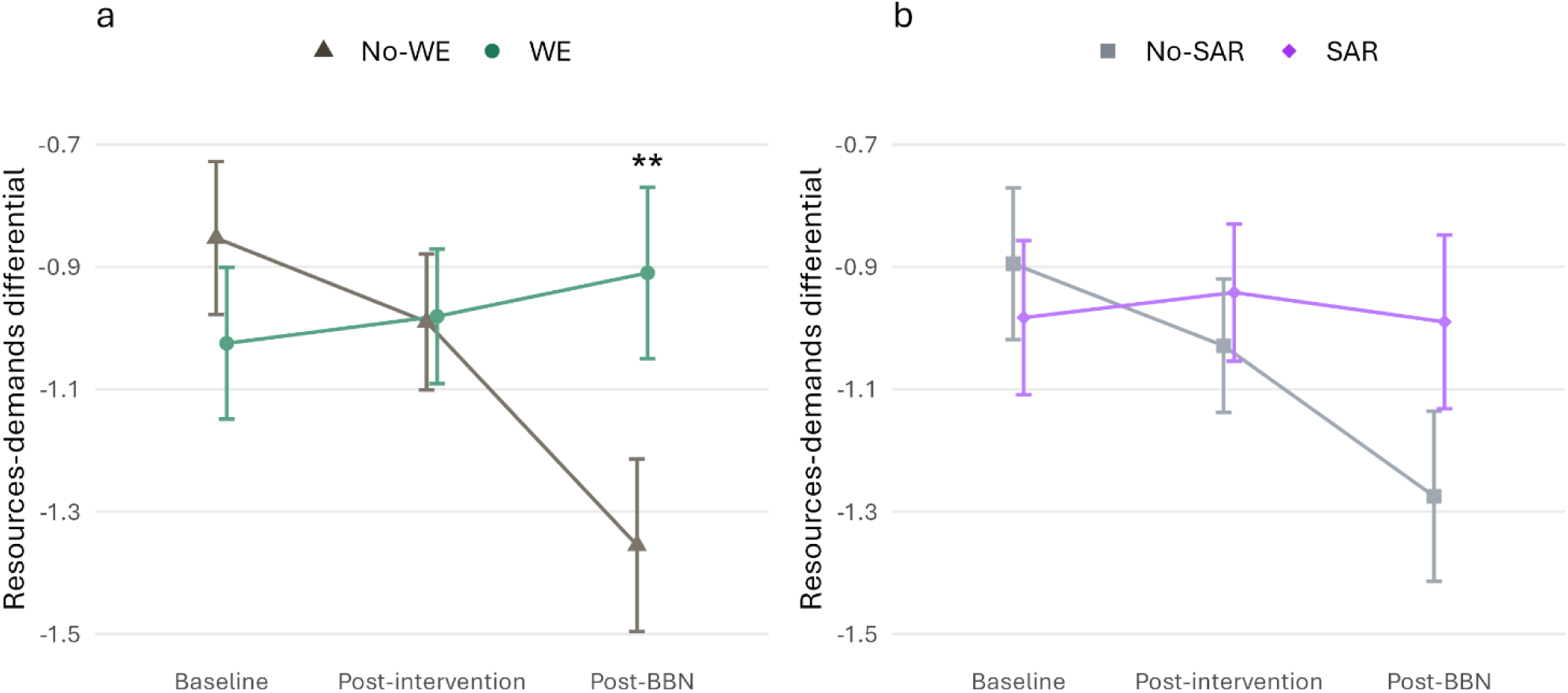
Figure 2. Line plots of Model 1 estimated marginal means of the resources-demands differential for WE vs. No-WE groups (a) and SAR vs. No-SAR groups (b). The error bar represents the standard error. Significant differences in the changes from baseline to respective time points are marked with asterisks (** *p* < .01).

#### Cardiovascular index of challenge and threat

In line with hypothesis 1.2, Model 1 showed a significant SAR × BBN interaction, indicating a significantly more positive change from baseline to BBN for the SAR groups (*M* = 0.46, *SE* = 0.13) than the No-SAR groups (*M* = -0.44, *SE* = 0.13), with a trend in the same direction for the changes from baseline to post-BBN. Contrary to hypothesis 2.2, no significant differences were observed between the WE groups (*M* = 0.00, *SE* = 0.13) and the No-WE groups (*M* = 0.02, *SE* = 0.13) in their changes from baseline to BBN, nor from baseline to other periods.

Model 2 revealed a significant SAR × WE × BBN interaction. Post-hoc analysis showed that the SAR x BBN effect was significant for the SAR-only group (*M* = 0.68, *SE* = 0.18) vs. the No-intervention group (*M* = -0.63, *SE* = 0.18), whereas the SAR x BBN effect for the SAR & WE group (*M* = 0.22, *SE* = 0.19) vs. the WE-only group (*M* = -0.25, *SE* = 0.18) approached significance. The WE x BBN effect approached significance for the SAR & WE group vs. the SAR-only group, whereas it was not significant for the WE-only group vs. the No-intervention group (see Table 3 and Fig. 3).

**Figure 3.**
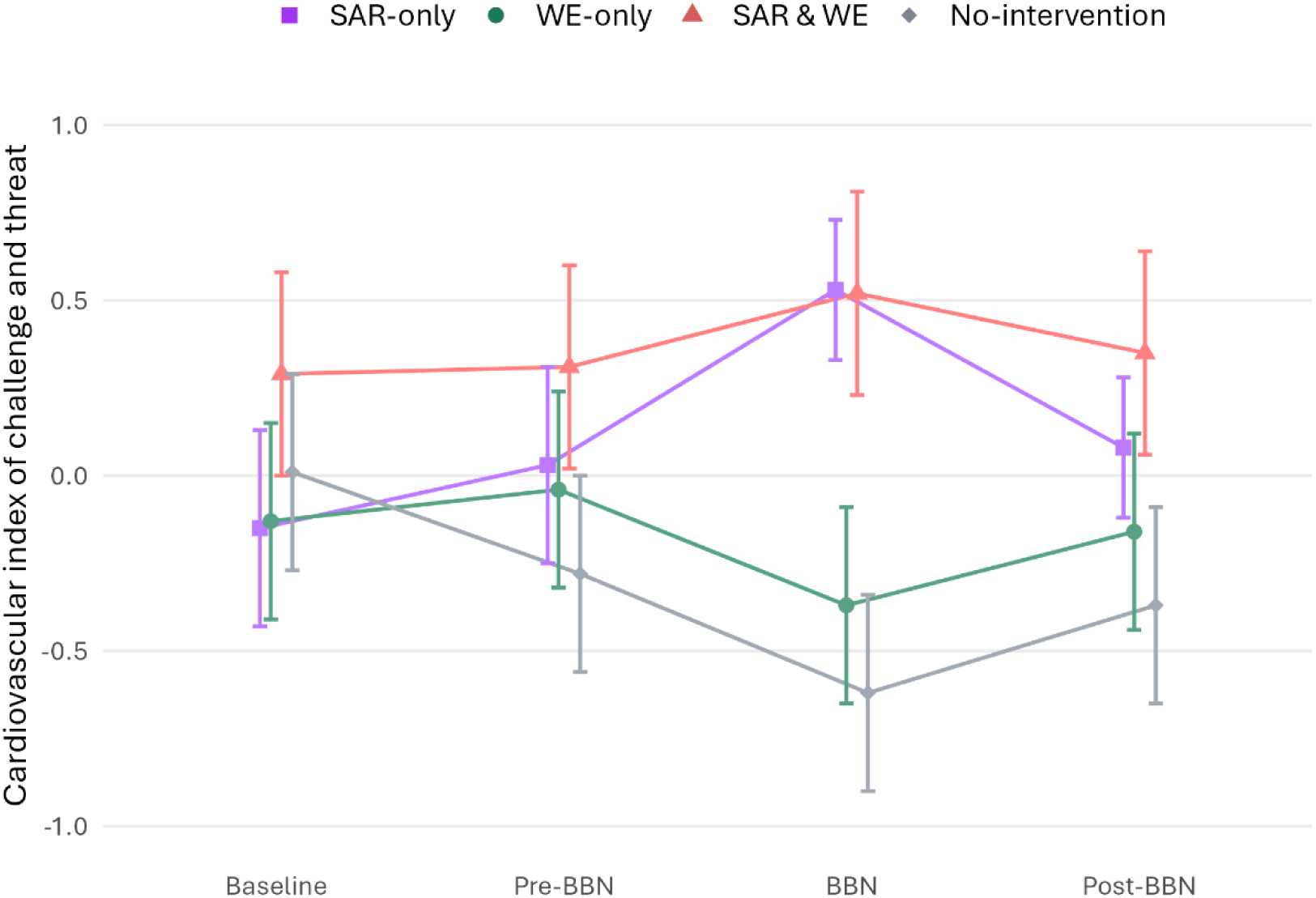
Line plot of the Model 2 estimated marginal means of the cardiovascular index of challenge and threat for the SAR-only, WE-only, SAR & WE, and No-intervention groups. The error bar represents the standard error. For contrast effects see Table 3.

**Table 3.**
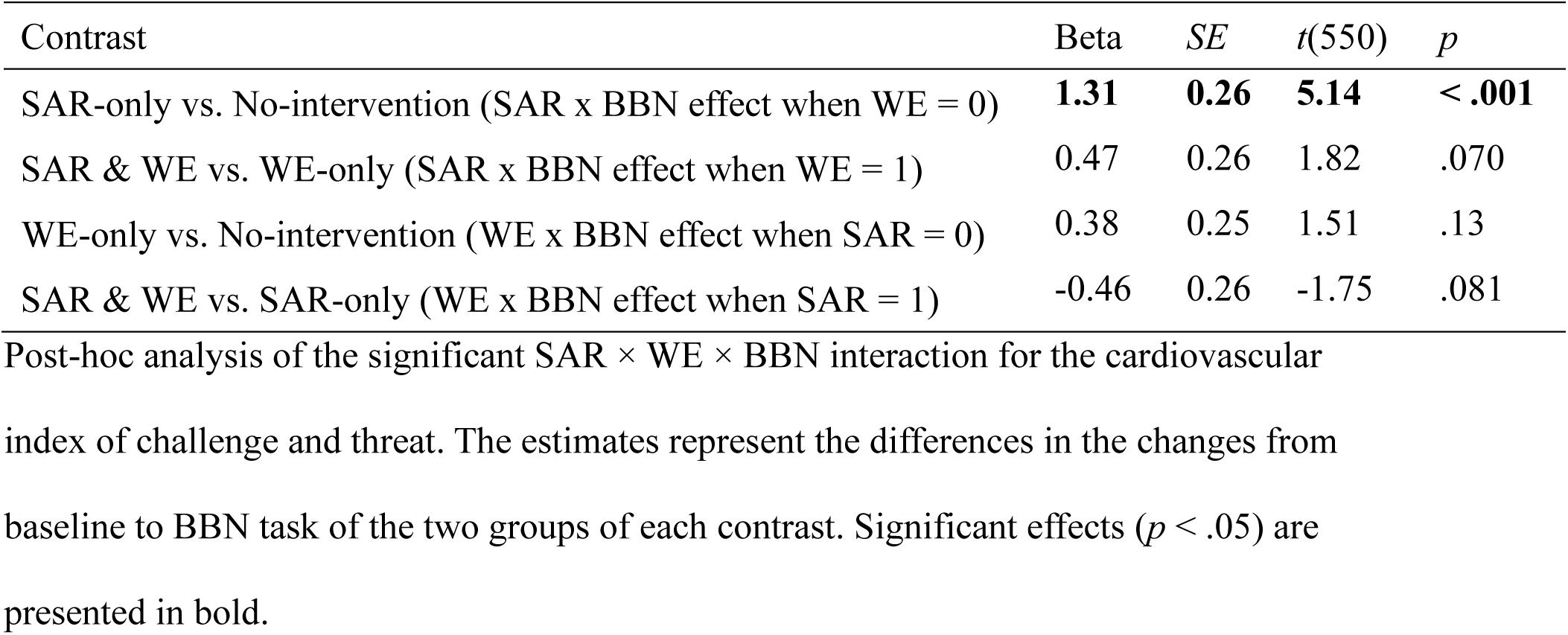

### Secondary outcomes

Descriptive statistics for the secondary outcomes are reported in the Supplementary Tables S2 and S3.

#### Demand and resource evaluations

As predicted, both interventions had no significant effects on changes in demand evaluation. As expected, we found a significant WE × Post-BBN interaction for resource evaluation, indicating a more positive change from baseline to post-BBN for the WE groups (*M* = 0.12, *SE* = 0.10) than the No-WE groups (*M* = -0.25, *SE* = 0.10). In contrast, the expected effect of SAR × Post-BBN was not significant (see Supplementary Tables S4 and S5 and Supplementary Fig. S1).

#### Cardiac output, total peripheral resistance, and stroke volume

In line with our secondary hypotheses, analyses of CO, TPR, and SV revealed significant SAR × BBN interactions, indicating improved changes from baseline to BBN for the SAR groups compared to the No-SAR groups. Specifically, we observed a larger increase in CO (SAR groups: *M* = 1.33, *SE* = 0.10; No-SAR groups: *M* = 0.56, *SE* = 0.09), a smaller increase in TPR (SAR groups: *M* = 2.12, *SE* = 0.30; No-SAR groups: *M* = 3.89, *SE* = 0.29), and a smaller decrease in SV (SAR groups: *M* = -2.41, *SE* = 1.00; No-SAR groups: *M* = - 8.20, *SE* = 0.97). Additionally, the SAR × post-BBN interaction for CO was also significant in the same direction. In contrast, the WE x BBN interaction was not significant for any parameters.

Similar to the cardiovascular index, a significant SAR × WE × BBN interaction was found for CO only. Post-hoc analysis revealed that the SAR x BBN effect was significant for the SAR-only group (*M* = 1.47, *SE* = 0.13) vs. the No-intervention group (*M* = 0.32, *SE* = 0.13), and the WE x BBN effect was significant for the WE-only group (*M* = 0.81, *SE* = 0.13) vs. the No-intervention group. Additionally, the SAR × WE × post-BBN interaction for CO was also significant. Post-hoc analysis revealed significant effects between SAR-only vs. No-intervention and WE-only vs. No-intervention (see Supplementary Tables S6-S9 and Supplementary Fig. S1 and S2).

### Task engagement

The descriptive statistics for the indices of task engagement HR and PEP are reported in the Supplementary Table S3.

#### Heart rate

As predicted, the sample as whole showed a significantly elevated HR before and during the BBN task, compared to baseline. We found no significant interactions (see Supplementary Table S10).

#### Pre-ejection period

In line with our predictions, the sample as a whole showed a significantly decreased PEP pre-BBN, during BBN and post-BBN, when compared to baseline. Model 1 revealed significant SAR × BBN and SAR × post-BBN interactions. The decrease in PEP from baseline to BBN was significantly larger for the SAR groups (*M* = -30.79, *SE* = 1.82) than the No-SAR groups (*M* = -22.92, *SE* = 1.77) as well as from baseline to post-BBN.

There was also a significant SAR × WE × BBN interaction. Post-hoc analysis revealed that the SAR x BBN effect was significant for the SAR-only group (*M* = -32.92, *SE* = 2.53) vs. the No-intervention group (*M* = -20.16, *SE* = 2.49) but not for the SAR & WE group (*M* = -28.45, *SE* = 2.60) vs. the WE-only group (*M* = -25.62, *SE* = 2.51). The WE x BBN effect was not significant either for the WE-only group vs. the No-intervention group or for the SAR & WE group vs. the SAR-only group (see Supplementary Tables S11-S12 and Supplementary Fig. 2).

## Discussion

The goal of the present study was to evaluate the effectiveness of SAR and WE-based learning interventions on the psychophysiological stress response of medical students tasked with BBN to simulated patients. The results revealed that the WE intervention led to a more favorable resources-demands differential after the BBN task, whereas the SAR instructions were effective in improving the students’ cardiovascular index of challenge and threat during the task. This latter effect was larger in the absence of the WE intervention. Overall, students were highly engaged in the BBN encounter as indicated by a significant increase in HR and a decrease in PEP from baseline to the BBN task.

### Effects of the stress arousal reappraisal intervention

The SAR intervention did not significantly affect the resources-demands evaluation, failing to confirm hypothesis 1.1. Although this finding contradicts previous research (e.g., ^28,30,47^), it is worth mentioning that the effect trended in the expected direction for the change from baseline to post-BBN task, both for the resources-demands differential and resource evaluation. Unique features in our study design compared to typical SAR research could explain the non-significant results. First, in our study, SAR was part of a broader learning module, competing with more extensive BBN instructions that were likely also more relevant to the novel BBN task. Second, our sample of third-year medical students already had a thorough understanding of stress physiology, so the SAR intervention may not have introduced new insights at first glance. As a result, the conscious perception of personal resources may have been less impacted, in contrast with previous studies where SAR instructions were the primary focus.

While the SAR intervention did not significantly influence the resources-demands differential, its significant effect on the cardiovascular index of challenge and threat is in accordance with hypothesis 1.2. In light of previously ambiguous results (e.g.,^29,43,46^), the effect observed in this study may partly be attributed to the specific characteristics of our population. Medical students, with their comprehensive understanding of stress physiology, might have found the positive reframing of stress arousal particularly compelling. Already aware of the beneficial aspects of stress, they may have been more open to reappraising their arousal. Interestingly, the SAR intervention was more effective for participants who received the SAR intervention only (compared to the No-intervention group) than for those who received both interventions (compared to the WE-only group). It is possible that participants engaged less in the SAR intervention when they were also presented with the WE, because they perceived the WE as more relevant to the BBN task (as reflected in the self-rated resource evaluation).

The SAR intervention showed similar positive effects on the secondary cardiovascular outcomes CO, TPR and SV, although its influence on SV and TPR was not moderated by the WE. The results suggest that not all cardiovascular measures reflect challenge and threat in the same way. The underlying reasons for these differences are difficult to identify. Notably, conceptual differences related to the BPSM of challenge and threat may be of interest. For instance, SV serves as an indicator of challenge and threat independent of HR, which, according to the BPSM of challenge and threat, is irrelevant for differentiating between challenge and threat^30,35,36^.

### Effects of the worked example-based learning intervention

In line with hypothesis 2.1, students who learned with the WE showed a more positive change from baseline to post-BBN in their resources-demands differential than students who did not receive the WE. As anticipated, perceived resources were the primary factor driving this effect, while demands remained largely unaffected by the WE intervention. These findings are in agreement with the idea that the WE promotes the acquisition of applicable skills that are perceived as personal resources to cope with the BBN task demands.

In contrast, the lack of a significant effect of WE on the cardiovascular index of challenge and threat does not support hypothesis 2.2. Although the WE improved self-reported perceived resources, it is possible that it unconsciously increased performance pressure by setting high standards that students felt they might not meet. According to the BPSM of challenge and threat, the evaluation of demands and resources occurs unconsciously. One key advantage of using cardiovascular measurements to assess challenge and threat is that they do not rely on an individual’s ability or willingness to accurately report on their appraisals^84,85^. Cardiovascular measures of challenge and threat states have been preferred over self-reported measures in the context of more subtle, extraneous factors (e.g.,^19^). In this context, it has been shown that the cardiovascular challenge and threat patterns can be influenced subconsciously^27^. Therefore, a discrepancy between self-reported and cardiovascular assessments of challenge and threat is possible (e.g.,^86^) and could explain the lack of effect of the WE intervention on the cardiovascular index of challenge and threat in our study. However, it is important to note that the WE intervention significantly improved the secondary cardiovascular outcome CO, when the SAR intervention was not administered concurrently. Again, this discrepancy in the results among cardiovascular outcomes highlights that different parameters may not capture challenge and threat responses to the same extent.

### Task engagement

The BPSM of challenge and threat applies to motivated performance situations that are characterized by task engagement^24^. In our study, significant increases in HR and decreases in PEP from a resting baseline period suggest that simulated BBN consultations are engaging for the medical students. These results are in line with research examining stress during simulated BBN encounters^8^, as well as findings regarding social evaluative speech tasks in the SAR domain (e.g.,^28,45,46^). It should be noted that task engagement was already evident before the BBN task began, and therefore before the occurrence of physiological changes caused by the act of speaking. Elevated physiological arousal is essential for SAR interventions to be effective since SAR is only feasible if there is an arousal to begin with^87^. Our study confirms that simulated BBN consultations are highly engaging and subjectively demanding to warrant stress management interventions.

An unexpected finding was a significantly larger decrease in PEP for the SAR groups than the No-SAR groups, particularly for participants who did not receive the WE. This suggests that SAR may have heightened task engagement, possibly enhancing its effectiveness.

Regarding the differences in the results for HR and PEP, there are arguments to be made for both parameters as preferable indicator of task engagement. On the one hand, whereas HR is affected by both sympathetic and parasympathetic activity^88^, PEP is more directly influenced by the sympathetic nervous system^32^. On the other hand, it is unclear whether differences in PEP responses reflect differences in task engagement or in challenge vs. threat^33,75,89^, or even related phenomena such as mental effort^90,91^. In the SAR domain, both HR (e.g.,^43,47^) and PEP (e.g.,^21,45^) have been widely used as indicators of task engagement. Intervention-related effects on PEP reactivity were typically not observed (^29,44,45^; but see^21^), although PEP has been utilized as an indicator of challenge and threat during recovery^35,36^.

### Strengths, limitations, and outlook

The strength of this study lies in its unique and comprehensive approach to the highly relevant task of BBN in healthcare. In a simulated environment, we were able to investigate SAR and WE interventions in BBN without jeopardizing the wellbeing of actual patients, thereby making a contribution to the advancement of medical education. For a first time, we were able to demonstrate that WE-based learning can foster a psychological challenge-oriented stress response and further improve specific cardiovascular parameters, in this case CO. Furthermore, we gained novel insights into the applicability of SAR interventions in medical training and its positive impact during BBN. The low threshold of both interventions makes them easy to integrate into academic curricula. Finally, the study contributes to a comprehensive understanding of the psychophysiology of medical students during BBN encounters. The results provide a thorough analysis of key psychological and cardiovascular indices considered in the BPSM of challenge and threat, which are rarely evaluated simultaneously.

We acknowledge that certain limitations constrain the generalizability of our findings. The changes in psychophysiological stress responses were observed under controlled experimental conditions. It is important to acknowledge that in the experiment, social evaluation was induced by the recording of the participants’ BBN performance and their interaction with the simulated patient, whereas in clinical settings, the social evaluation may stem from judgements made by patients, family members, and supervisors. Moreover, the experimental setting meant that the students interacted with simulated rather than real patients. However, the voluntary participation of the students in the experiment implies their willingness to immerse themselves in the simulation, with the overarching goal of learning from this experience. Furthermore, simulated situations are standard procedure in medical education and assessment, which may facilitate students’ ability to suspend disbelief^92^. Despite the inherent differences between simulated and clinical settings, we are optimistic that similar improvements can be achieved in real-life applications^93^. The observed task engagement and high motivation to perform well on the task may help to mitigate some of the concerns in this regard.

Future research may want to investigate the interventions within a clinical setting, while also considering ethical implications. Further, it would be valuable to explore the effectiveness of the SAR intervention among more experienced health care providers, as increased experience may alter stress responses^94^. Exploring the sustainability of SAR effects, along with potential spillover effects, represents important and underexamined directions for future research. More generally, an intriguing research question involves identifying the specific stressors and individual stress thresholds at which the SAR intervention proves most effective in regulating the psychophysiological responses. Furthermore, future studies could enhance understanding of potential WE effects on cardiovascular responses, considering the observed discrepancy between the cardiovascular and self-reported measures. Given the partially divergent effects of the interventions on different cardiovascular outcomes, further research is needed to clarify the meaning and relevance of these parameters within the BPSM of challenge and threat.

### Implications and conclusion

The present findings offer valuable insights into both the psychophysiological dynamics of BBN and the potential for stress management interventions in medical education.

From a theoretical perspective, WE-based learning, which directly contributes to successful task completion, resulted in a more advantageous psychological stress evaluation. This demonstrates that WEs, traditionally applied in well-structured fields like mathematics, can also be beneficial in complex communication tasks such as BBN. Conversely, SAR improved mainly the cardiovascular stress response, confirming its effectiveness in distressing communication tasks and suggesting a high receptiveness among a primed population. Our findings revealed slightly deviating effects of the interventions on the varying cardiovascular parameters within the BPSM of challenge and threat, pointing towards subtle yet meaningful differences in how these parameters capture challenge and threat. This variability in effects raises concerns about relying on a single cardiovascular parameter within the BPSM, as study findings may depend on the chosen measure. In the absence of standardized guidelines, using multiple parameters may offer a more robust and comprehensive evaluation.

From a practical standpoint, our study shows that simulated BBN scenarios elicit heightened physiological arousal in third-year medical students. Notably, the resource-demand differential consistently indicated a threat state across all periods, underscoring the need for stress management programs in medical education. SAR stands out as a time-and cost-efficient intervention that can easily be integrated into already crowded curricula. The potential for far-reaching spillover effects is notable in both educational and clinical workplace environments, which are rife with stressful situations. Although there is no panacea for teaching stressful communication tasks, our study demonstrates that subjective benefits can arise from WE-based learning. Overall, our findings suggest that both SAR and WE could help optimize physicians’ psychophysiological responses when delivering bad news. However, implementing the interventions separately for novice learners may be more effective, as simultaneous application could divide resources and attention, reducing their positive impact at least on the cardiovascular outcomes.

## Data Availability

All data produced in the present study are available upon reasonable request to the authors

https://osf.io/9aqwn/

## Data availability

Data supporting the findings of this study are available in the tables and figures of this article. The datasets generated and analyzed during the current study are available in the OSF repository https://osf.io/9aqwn/.

## Author contributions

**Michel Bosshard:** Methodology, Formal analysis, Investigation, Resources, Writing – Original Draft, Visualization. **Sissel Guttormsen:** Conceptualization, Methodology, Writing – Review & Editing, Funding Acquisition. **Urs Markus Nater:** Conceptualization, Methodology, Writing – Review & Editing, Funding Acquisition. **Felix Schmitz:** Conceptualization, Methodology, Writing – Review & Editing, Supervision, Project administration, Funding acquisition. **Patrick Gomez:** Conceptualization, Methodology, Formal analysis, Writing – Review & Editing, Supervision, Funding acquisition. **Christoph Berendonk:** Conceptualization, Methodology, Writing – Review & Editing, Supervision, Project administration, Funding acquisition.

All authors have approved the submitted version (and any substantially modified version that involves the author’s contribution to the study) and have agreed both to be personally accountable for the author’s own contributions and to ensure that questions related to the accuracy or integrity of any part of the work, even ones in which the author was not personally involved, are appropriately investigated, resolved, and the resolution documented in the literature.

## Additional Information

The authors declare no competing interests.

