## Supplementary Material for "Optimizing stress in breaking bad news: a randomized controlled trial on the psychophysiological effects of stress arousal reappraisal and worked-example interventions among medical students"

Michel Bosshard[1,2*], Sissel Guttormsen[1], Urs Markus Nater[3,4], Felix Schmitz[1 †], Patrick Gomez[5 †], Christoph Berendonk[1 †]

[1]Institute for Medical Education, University of Bern, Bern, Switzerland

[2]Graduate School for Health Sciences, University of Bern, Bern, Switzerland

[3]Department of Clinical and Health Psychology, University of Vienna, Vienna, Austria

[4]University Research Platform “Stress of life (SOLE) – Processes and Mechanisms underlying everyday Life Stress”, University of Vienna, Vienna, Austria

[5] Department of Occupational and Environmental Health, Unisanté, Center for Primary Care and Public Health & University of Lausanne, Lausanne, Switzerland

* Corresponding Author

† Felix Schmitz, Patrick Gomez and Christoph Berendonk share last authorship

**Supplementary Table S1**

| | SAR-only ($n$ = 55) | WE-only ($n$ = 58) | SAR & WE ($n$ = 57) | No-intervention ($n$ = 59) | Effect |
|---|---|---|---|---|---|
| BMI (kg/m$^2$) | 22.29 (2.89) | 22.12 (2.64) | 22.39 (2.69) | 22.49 (2.97) | $F$(3, 225) = 0.18, $p$ = .91 |
| Sex (% female) | 70.9 | 69.0 | 68.4 | 67.8 | $X^2$ (3, $N$ = 229) = 0.14, $p$ = .99 |
| Age (in years) | 22.42 (2.17) | 22.24 (1.33) | 22.60 (1.81) | 22.42 (1.94) | $F$(3, 225) = 0.36, $p$ = .78 |
| Shift work (% yes) | 27.3 | 29.3 | 19.3 | 27.1 | $X^2$ (3, $N$ = 229) = 1.75, $p$ = .63 |
| Contraceptives (% yes) | 36.4 | 36.2 | 29.8 | 35.6 | $X^2$ (3, $N$ = 229) = 0.74, $p$ = .86 |
| Depression | 2.7 (2.9) | 1.9 (2.0) | 2.7 (3.1) | 2.5 (2.6) | $F$(3, 225) = 1.25, $p$ = .29 |
| Anxiety | 2.4 (3.0) | 2.9 (2.5) | 2.6 (2.5) | 2.6 (2.8) | $F$(3, 225) = 0.31, $p$ = .82 |
| Stress | 4.4 (3.4) | 4.2 (3.4) | 3.6 (3.6) | 4.7 (3.7) | $F$(3, 225) = 0.96, $p$ = .41 |
| BBN skills | 4.1 (0.9) | 4.1 (1.0) | 3.9 (0.9) | 3.9 (0.9) | $F$(3, 187) = 0.57, $p$ = .64 |
| BBN experience (% yes) | 18.2 | 6.9 | 8.8 | 8.5 | $X^2$ (3, $N$ = 229) = 4.70, $p$ = .20 |
| BBN interest | 5.9 (0.9) | 5.8 (0.9) | 5.9 (0.8) | 5.6 (0.9) | $F$(3, 225) = 1.48, $p$ = .22 |
| BBN motivation | 6.5 (0.6) | 6.0 (0.9) | 6.3 (0.7) | 6.3 (0.9) | $F$(3, 225) = 3.89, $p$ = .010 |
| BBN task duration (in seconds) | 528 (113) | 506 (105) | 507 (110) | 494 (121) | $F$(3, 225) = 0.89, $p$ = .45 |

Sociodemographic and BBN-related variables for the four experimental groups. Mean values with standard deviation in parentheses. BMI = Body Mass Index.

**Supplementary Table S2**

| Variable | Period | SAR-only ($n = 55$) | WE-only ($n = 58$) | SAR & WE ($n = 57$) | No-intervention ($n = 59$) | Effect |
|---|---|---|---|---|---|---|
| Resources-Demands differential | Baseline | -0.84 (0.17) | -0.97 (0.18) | -1.05 (0.14) | -0.85 (0.20) | $F(3, 225) = 0.51$ $p = .68$ |
| | Post-intervention | -0.85 (0.14) | -1.05 (0.17) | -0.96 (0.13) | -1.07 (0.19) | |
| | Post-BBN | -1.09 (0.21) | -1.02 (0.19) | -0.84 (0.18) | -1.58 (0.19) | |
| Demand evaluation | Baseline | 4.38 (0.11) | 4.41 (0.11) | 4.51 (0.08) | 4.32 (0.11) | $F(3, 225) = 0.56$ $p = .65$ |
| | Post-intervention | 4.56 (0.11) | 4.62 (0.11) | 4.67 (0.09) | 4.51 (0.09) | |
| | Post-BBN | 4.53 (0.16) | 4.48 (0.13) | 4.46 (0.13) | 4.66 (0.11) | |
| Resource evaluation | Baseline | 3.55 (0.12) | 3.45 (0.12) | 3.40 (0.11) | 3.47 (0.13) | $F(3, 225) = 0.23$ $p = .87$ |
| | Post-intervention | 3.71 (0.11) | 3.57 (0.11) | 3.70 (0.10) | 3.44 (0.12) | |
| | Post-BBN | 3.44 (0.14) | 3.47 (0.11) | 3.61 (0.11) | 3.08 (0.11) | |

Descriptive statistics of the resource and demand outcomes for the four experimental groups. Raw mean values with standard error in parentheses.

**Supplementary Table S3**

| Variable | Period | SAR-only ($n$ = 48) | WE-only ($n$ = 49) | SAR & WE ($n$ = 44) | No-intervention ($n$ = 48) | Effect |
|---|---|---|---|---|---|---|
| Cardiovascular index | Baseline | -0.17 (0.33) | -0.11 (0.28) | 0.29 (0.24) | 0.01 (0.24) | $F$(3, 185) = 0.52 $p$ = .67 |
| | Pre-BBN | 0.01 (0.30) | -0.15 (0.31) | 0.31 (0.19) | -0.28 (0.27) | |
| | BBN | 0.53 (0.29) | -0.37 (0.29) | 0.52 (0.22) | -0.62 (0.25) | |
| | Post-BBN | 0.06 (0.32) | 0.00 (0.26) | 0.35 (0.23) | -0.38 (0.29) | |
| Cardiac output (in L/min) | Baseline | 6.03 (0.20) | 6.05 (0.16) | 6.25 (0.17) | 6.12 (0.16) | $F$(3, 185) = 0.31, $p$ = .82 |
| | Pre-BBN | 6.73 (0.21) | 6.80 (0.22) | 6.90 (0.16) | 6.45 (0.21) | |
| | BBN | 7.51 (0.25) | 6.85 (0.19) | 7.43 (0.19) | 6.44 (0.19) | |
| | Post-BBN | 6.14 (0.22) | 6.06 (0.16) | 6.21 (0.17) | 5.72 (0.18) | |
| Total peripheral resistance (in mmHg·min/L) | Baseline | 15.65 (0.76) | 15.45 (0.65) | 14.49 (0.46) | 15.22 (0.48) | $F$(3, 185) = 0.67, $p$ = .57 |
| | Pre-BBN | 17.34 (0.80) | 17.72 (0.82) | 16.49 (0.43) | 17.74 (0.65) | |
| | BBN | 17.28 (0.62) | 19.33 (0.81) | 17.07 (0.51) | 19.18 (0.61) | |
| | Post-BBN | 19.65 (0.89) | 19.62 (0.77) | 18.36 (0.60) | 20.27 (0.84) | |

**Supplementary Table S3 (continued)**

| Variable | Period | SAR-only ($n$ = 48) | WE-only ($n$ = 49) | SAR & WE ($n$ = 44) | No-intervention ($n$ = 48) | Effect |
|---|---|---|---|---|---|---|
| Stroke Volume (in mL) | Baseline | 80.81 (2.81) | 78.14 (2.12) | 77.77 (1.93) | 78.38 (2.28) | $F(3, 185)$ = 0.35, $p$ = .79 |
| | Pre-BBN | 75.90 (2.53) | 82.73 (1.98) | 73.02 (1.92) | 70.96 (2.45) | |
| | BBN | 78.25 (2.74) | 71.61 (2.07) | 75.45 (2.00) | 68.48 (2.23) | |
| | Post-BBN | 83.21 (3.26) | 79.52 (2.50) | 77.43 (2.35) | 76.70 (2.98) | |
| Heart Rate (in bpm) | Baseline | 75.85 (1.93) | 78.41 (1.85) | 80.98 (1.87) | 79.29 (1.88) | $F(3, 185)$ = 1.26, $p$ = .29 |
| | Pre-BBN | 89.98 (2.31) | 94.71 (2.92) | 96.02 (2.45) | 92.60 (2.45) | |
| | BBN | 97.67 (2.59) | 96.73 (1.94) | 99.73 (2.31) | 96.02 (2.59) | |
| | Post-BBN | 75.31 (2.13) | 77.14 (1.65) | 81.57 (2.04) | 77.29 (2.25) | |
| Pre-Ejection Period (in ms) | Baseline | 117.60 (2.12) | 114.67 (2.37) | 112.52 (2.12) | 107.81 (2.68) | $F(3, 185)$ = 3.15, $p$ = .026 |
| | Pre-BBN | 98.67 (3.25) | 96.57 (3.02) | 90.16 (3.52) | 94.73 (3.36) | |
| | BBN | 85.44 (3.09) | 88.31 (3.07) | 84.11 (3.10) | 87.63 (3.10) | |
| | Post-BBN | 101.75 (2.62) | 105.37 (2.36) | 101.30 (2.29) | 99.77 (3.13) | |

Descriptive statistics of the cardiovascular outcomes for the four experimental groups. Raw mean values with standard error in parentheses.

**Supplementary Table S4**

| Fixed Effect | Model 1 | | | Model 2 | | |
|---|---|---|---|---|---|---|
| | Beta | *SE* | *p* | Beta | *SE* | *p* |
| Intercept | 3.96 | 0.45 | < .001 | 3.96 | 0.45 | < .001 |
| SAR [a] | 0.05 | 0.14 | .72 | 0.05 | 0.15 | .76 |
| WE [b] | 0.11 | 0.14 | .43 | 0.11 | 0.15 | .48 |
| Post-Intervention [c] | 0.76 | 0.41 | .062 | 0.75 | 0.41 | .069 |
| Post-BBN [c] | 0.67 | 0.55 | .23 | 0.69 | 0.56 | .22 |
| Motivation [d] | 0.06 | 0.07 | .42 | 0.06 | 0.07 | .42 |
| SAR × WE | 0.02 | 0.18 | .89 | 0.03 | 0.21 | .89 |
| SAR × Post-Intervention | -0.00 | 0.10 | .97 | 0.02 | 0.14 | .91 |
| SAR× Post-BBN | -0.14 | 0.13 | .29 | -0.18 | 0.19 | .34 |
| WE × Post-Intervention | -0.02 | 0.10 | .82 | -0.00 | 0.14 | .98 |
| WE × Post-BBN | -0.25 | 0.13 | .064 | -0.29 | 0.19 | .13 |
| Motivation × Post-Intervention | -0.09 | 0.06 | .16 | -0.09 | 0.06 | .16 |
| Motivation × Post-BBN | -0.05 | 0.09 | .52 | -0.06 | 0.09 | .52 |
| SAR × WE × Post-Intervention | | | | -0.04 | 0.19 | .85 |
| SAR × WE × Post-BBN | | | | 0.08 | 0.26 | .77 |

Linear mixed effects regressions for the demand evaluation. [a] No-SAR = 0, SAR = 1. [b] No-WE = 0, WE = 1. [c] Reference = baseline. [d] Motivation is grand mean centered.

**Supplementary Table S5**

| Fixed Effect | Model 1 | | | Model 2 | | |
|---|---|---|---|---|---|---|
| | Beta | *SE* | *p* | Beta | *SE* | *p* |
| Intercept | 2.53 | 0.51 | < .001 | 2.54 | 0.52 | < .001 |
| SAR [a] | 0.06 | 0.15 | .72 | 0.04 | 0.17 | .83 |
| WE [b] | 0.03 | 0.15 | .84 | 0.01 | 0.17 | .94 |
| Post-Intervention [c] | -0.40 | 0.52 | .45 | -0.41 | 0.53 | .44 |
| Post-BBN [c] | -0.51 | 0.59 | .38 | -0.54 | 0.59 | .37 |
| Motivation [d] | 0.15 | 0.08 | .062 | 0.15 | 0.08 | .063 |
| SAR × WE | -0.16 | 0.18 | .36 | -0.13 | 0.24 | .60 |
| SAR × Post-Intervention | 0.17 | 0.13 | .17 | 0.18 | 0.18 | .30 |
| SAR × Post-BBN | 0.23 | 0.14 | .10 | 0.28 | 0.20 | .17 |
| WE × Post-Intervention | 0.16 | 0.13 | .21 | 0.17 | 0.18 | .33 |
| WE × Post-BBN | **0.37** | **0.14** | **.009** | 0.41 | 0.20 | .037 |
| Motivation × Post-Intervention | 0.06 | 0.08 | .47 | 0.06 | 0.08 | .47 |
| Motivation × Post-BBN | 0.02 | 0.09 | .80 | 0.02 | 0.09 | .80 |
| SAR × WE × Post-Intervention | | | | -0.02 | 0.25 | .92 |
| SAR × WE × Post-BBN | | | | -0.09 | 0.28 | .75 |

Linear mixed effects regressions for the resource evaluation. Significant interaction effects ($p < .05$) are presented in bold. [a] No-SAR = 0, SAR = 1. [b] No-WE = 0, WE = 1. [c] Reference = baseline. [d] Motivation is grand mean centered.

**Supplementary Table S6**

| Fixed Effect | Model 1 | | | Model 2 | | |
|---|---|---|---|---|---|---|
| | Beta | *SE* | *p* | Beta | *SE* | *p* |
| Intercept | 6.04 | 0.18 | < .001 | 6.12 | 0.18 | < .001 |
| SAR [a] | 0.07 | 0.25 | .77 | -0.10 | 0.26 | .70 |
| WE [b] | 0.11 | 0.25 | .66 | -0.06 | 0.25 | .82 |
| Pre-BBN [c] | 0.44 | 0.11 | < .001 | 0.33 | 0.13 | .011 |
| BBN [c] | 0.51 | 0.12 | < .001 | 0.32 | 0.13 | .016 |
| Post-BBN [c] | -0.26 | 0.10 | .007 | -0.38 | 0.11 | < .001 |
| Motivation [d] | 0.04 | 0.12 | .76 | 0.04 | 0.12 | .76 |
| SAR × WE | -0.06 | 0.34 | .86 | 0.28 | 0.36 | .43 |
| SAR × Pre-BBN | 0.14 | 0.13 | .29 | 0.37 | 0.18 | .043 |
| SAR × BBN | **0.77** | **0.14** | **< .001** | 1.15 | 0.19 | < .001 |
| SAR × Post-BBN | **0.26** | **0.11** | **.025** | 0.49 | 0.16 | .002 |
| WE × Pre-BBN | 0.19 | 0.13 | .15 | 0.42 | 0.18 | .022 |
| WE × BBN | 0.11 | 0.14 | .43 | 0.49 | 0.19 | .009 |
| WE × Post-BBN | 0.09 | 0.11 | .42 | 0.32 | 0.16 | .039 |
| Motivation × Pre-BBN | -0.02 | 0.08 | .77 | -0.02 | 0.08 | .77 |
| Motivation × BBN | 0.02 | 0.09 | .82 | 0.02 | 0.08 | .82 |
| Motivation × Post-BBN | -0.03 | 0.07 | .70 | -0.03 | 0.07 | .71 |
| SAR × WE × Pre-BBN | | | | -0.47 | 0.26 | .070 |
| SAR × WE × BBN | | | | **-0.78** | **0.26** | **.003** |
| SAR × WE × Post-BBN | | | | **-0.47** | **0.22** | **.032** |

Linear mixed effects regressions for cardiac output. Significant interaction effects ($p < .05$) are presented in bold. [a] No-SAR = 0, SAR = 1. [b] No-WE = 0, WE = 1. [c] Reference = baseline. [d] Motivation is grand mean centered.

**Supplementary Table S7**

| Contrast | Beta | *SE* | *t*(550) | *p* |
|---|---|---|---|---|
| SAR × WE × BBN | | | | |
| SAR-only vs. No-intervention (SAR x BBN effect when WE = 0) | **1.15** | **0.19** | **6.17** | **< .001** |
| SAR & WE vs. WE-only (SAR x BBN effect when WE = 1) | 0.37 | 0.19 | 1.95 | .051 |
| WE-only vs. No-intervention (WE x BBN effect when SAR = 0) | **0.49** | **0.19** | **2.63** | **.009** |
| SAR & WE vs. SAR-only (WE x BBN effect when SAR = 1) | -0.29 | 0.19 | -1.54 | .12 |
| SAR × WE × post-BBN | | | | |
| SAR-only vs. No-intervention (SAR x post-BBN effect when WE = 0) | **0.49** | **0.16** | **3.12** | **.002** |
| SAR & WE vs. WE-only (SAR x post-BBN effect when WE = 1) | 0.02 | 0.16 | 0.10 | .92 |
| WE-only vs. No-intervention (WE x post-BBN effect when SAR = 0) | **0.32** | **0.16** | **2.07** | **.039** |
| SAR & WE vs. SAR-only (WE x post-BBN effect when SAR = 1) | -0.15 | 0.16 | -0.95 | .34 |

Post-hoc analysis of the significant SAR × WE × BBN and SAR × WE × post-BBN interaction for cardiac output. The estimates represent the differences in the changes from baseline to BBN task and from baseline to post-BBN of the two groups of each contrast. Significant effects ($p$ < .05) are presented in bold.

**Supplementary Table S8**

| Fixed Effect | Model 1 | | | Model 2 | | |
|---|---|---|---|---|---|---|
| | Beta | *SE* | *p* | Beta | *SE* | *p* |
| Intercept | 15.36 | 0.67 | < .001 | 15.23 | 0.68 | < .001 |
| SAR [a] | 0.03 | 0.94 | .97 | 0.31 | 0.98 | .75 |
| WE [b] | 0.08 | 0.93 | .93 | 0.35 | 0.97 | .72 |
| Pre-BBN [c] | 2.38 | 0.36 | < .001 | 2.52 | 0.42 | < .001 |
| BBN [c] | 3.70 | 0.35 | < .001 | 3.96 | 0.41 | < .001 |
| Post-BBN [c] | 4.95 | 0.45 | < .001 | 5.01 | 0.52 | < .001 |
| Motivation [d] | 0.40 | 0.44 | .37 | 0.40 | 0.44 | .37 |
| SAR × WE | -0.84 | 1.27 | .51 | -1.40 | 1.38 | .31 |
| SAR × Pre-BBN | -0.57 | 0.43 | .18 | -0.85 | 0.60 | .15 |
| SAR × BBN | **-1.76** | **0.42** | **< .001** | -2.27 | 0.58 | < .001 |
| SAR × Post-BBN | -0.88 | 0.53 | .099 | -1.02 | 0.74 | .17 |
| WE × Pre-BBN | 0.04 | 0.43 | .92 | -0.24 | 0.59 | .69 |
| WE × BBN | 0.36 | 0.42 | .38 | -0.14 | 0.58 | .81 |
| WE × Post-BBN | -0.28 | 0.53 | .60 | -0.41 | 0.73 | .57 |
| Motivation × Pre-BBN | 0.06 | 0.27 | .84 | 0.06 | 0.27 | .84 |
| Motivation × BBN | -0.20 | 0.26 | .44 | -0.20 | 0.26 | .44 |
| Motivation × Post-BBN | -0.02 | 0.33 | .95 | -0.02 | 0.33 | .95 |
| SAR × WE × Pre-BBN | | | | 0.57 | 0.84 | .50 |
| SAR × WE × BBN | | | | 1.04 | 0.82 | .21 |
| SAR × WE × Post-BBN | | | | 0.28 | 1.04 | .79 |

Linear mixed effects regressions for total peripheral resistance. Significant interaction effects ($p < .05$) are presented in bold. [a] No-SAR = 0, SAR = 1. [b] No-WE = 0, WE = 1. [c] Reference = baseline. [d] Motivation is grand mean centered.

**Supplementary Table S9**

| Fixed Effect | Model 1 | | | Model 2 | | |
|---|---|---|---|---|---|---|
| | Beta | *SE* | *p* | Beta | *SE* | *p* |
| Intercept | 77.83 | 2.32 | < .001 | 78.37 | 2.36 | < .001 |
| SAR [a] | 3.61 | 3.26 | .27 | 2.54 | 3.37 | .45 |
| WE [b] | 0.74 | 3.24 | .82 | -0.33 | 3.35 | .92 |
| Pre-BBN [c] | -6.97 | 1.10 | < .001 | -7.42 | 1.27 | < .001 |
| BBN [c] | -9.13 | 1.19 | < .001 | -9.90 | 1.37 | < .001 |
| Post-BBN [c] | -0.36 | 1.46 | .80 | -1.65 | 1.68 | .33 |
| Motivation [d] | -0.32 | 1.52 | .83 | -0.33 | 1.52 | .83 |
| SAR × WE | -4.99 | 4.42 | .26 | -2.81 | 4.77 | .56 |
| SAR × Pre-BBN | 1.80 | 1.30 | .17 | 2.70 | 1.81 | .14 |
| SAR × BBN | **5.79** | **1.41** | **< .001** | 7.32 | 1.95 | < .001 |
| SAR × Post-BBN | 1.53 | 1.73 | .37 | 4.09 | 2.39 | .088 |
| WE × Pre-BBN | 0.93 | 1.30 | .48 | 1.82 | 1.80 | .31 |
| WE × BBN | 1.86 | 1.40 | .19 | 3.38 | 1.94 | .082 |
| WE × Post-BBN | -0.12 | 1.72 | .94 | 2.42 | 2.38 | .31 |
| Motivation × Pre-BBN | -0.64 | 0.82 | .44 | -0.64 | 0.82 | .44 |
| Motivation × BBN | 0.03 | 0.88 | .97 | 0.03 | 0.88 | .97 |
| Motivation × Post-BBN | -0.14 | 1.08 | .90 | -0.14 | 1.08 | .89 |
| SAR × WE × Pre-BBN | | | | -1.84 | 2.56 | .47 |
| SAR × WE × BBN | | | | -3.12 | 2.76 | .26 |
| SAR × WE × Post-BBN | | | | -5.20 | 3.37 | .12 |

Linear mixed effects regressions for stroke volume. Significant interaction effects ($p < .05$) are presented in bold. [a] No-SAR = 0, SAR = 1. [b] No-WE = 0, WE = 1. [c] Reference = baseline. [d] Motivation is grand mean centered.

**Supplementary Table S10**

| Fixed Effect | Model 0 | | | Model 1 | | | Model 2 | | |
|---|---|---|---|---|---|---|---|---|---|
| | Beta | *SE* | *p* | Beta | *SE* | *p* | Beta | *SE* | *p* |
| Intercept | 76.99 | 1.71 | < .001 | 79.10 | 2.03 | < .001 | 79.30 | 2.08 | < .001 |
| SAR [a] | 0.53 | 1.97 | .79 | -3.23 | 2.85 | .26 | -3.62 | 2.97 | .22 |
| WE [b] | 2.72 | 1.96 | .17 | -0.33 | 2.83 | .91 | -0.71 | 2.95 | .81 |
| Pre-BBN [c] | **14.70** | **0.93** | **< .001** | 13.83 | 1.61 | < .001 | 13.32 | 1.86 | < .001 |
| BBN [c] | **18.90** | **0.79** | **< .001** | 17.88 | 1.36 | < .001 | 16.73 | 1.57 | < .001 |
| Post-BBN [c] | -0.84 | 0.61 | .17 | -2.10 | 1.06 | .048 | -2.00 | 1.22 | .10 |
| Motivation [d] | 0.58 | 1.23 | .64 | 0.58 | 1.34 | .66 | 0.58 | 1.34 | .67 |
| SAR × WE | | | | 5.21 | 3.85 | .18 | 6.00 | 4.19 | .15 |
| SAR × Pre-BBN | | | | -0.32 | 1.91 | .87 | 0.70 | 2.66 | .79 |
| SAR × BBN | | | | 2.65 | 1.62 | .10 | 4.94 | 2.24 | .028 |
| SAR × Post-BBN | | | | 1.73 | 1.25 | .17 | 1.54 | 1.74 | .38 |
| WE × Pre-BBN | | | | 2.09 | 1.90 | .27 | 3.10 | 2.64 | .24 |
| WE × BBN | | | | -0.54 | 1.61 | .74 | 1.73 | 2.22 | .44 |
| WE × Post-BBN | | | | 0.86 | 1.25 | .49 | 0.66 | 1.73 | .70 |
| Motivation × Pre-BBN | | | | 0.36 | 1.20 | .77 | 0.36 | 1.20 | .77 |
| Motivation × BBN | | | | 0.45 | 1.01 | .66 | 0.45 | 1.01 | .65 |
| Motivation × Post-BBN | | | | -0.25 | 0.79 | .75 | -0.25 | 0.79 | .75 |
| SAR × WE × Pre-BBN | | | | | | | -2.07 | 3.76 | .58 |
| SAR × WE × BBN | | | | | | | -4.66 | 3.16 | .14 |
| SAR × WE × Post-BBN | | | | | | | 0.40 | 2.46 | .87 |

Linear mixed effects regressions for heart rate**.** Significant main effects in Model 0 ($p < .05$) are presented in bold. [a] No-SAR = 0, SAR = 1. [b] No-WE = 0, WE = 1. [c] Reference = baseline. [d] Motivation is grand mean centered.

**Supplementary Table S11**

| Fixed Effect | Model 0 | | | Model 1 | | | Model 2 | | |
|---|---|---|---|---|---|---|---|---|---|
| | Beta | *SE* | *p* | Beta | *SE* | *p* | Beta | *SE* | *p* |
| Intercept | 112.95 | 2.13 | < .001 | 108.89 | 2.58 | < .001 | 107.82 | 2.67 | < .001 |
| SAR [a] | -0.54 | 2.37 | .82 | 7.47 | 3.57 | .038 | 9.61 | 3.81 | .013 |
| WE [b] | 0.99 | 2.37 | .67 | 4.91 | 3.55 | .17 | 7.03 | 3.79 | .065 |
| Pre-BBN [c] | **-18.03** | **1.39** | **< .001** | -13.47 | 2.38 | < .001 | -13.08 | 2.74 | < .001 |
| BBN [c] | **-26.75** | **1.27** | **< .001** | -22.60 | 2.17 | < .001 | -20.16 | 2.49 | **< .001** |
| Post-BBN [c] | **-11.10** | **1.15** | **< .001** | -9.48 | 1.95 | < .001 | -8.04 | 2.24 | **< .001** |
| Motivation [d] | 1.36 | 1.49 | .36 | 0.59 | 1.72 | .73 | 0.59 | 1.72 | .73 |
| SAR × WE | | | | -7.59 | 4.62 | .10 | -11.94 | 5.38 | .028 |
| SAR × Pre-BBN | | | | -5.23 | 2.82 | .064 | -6.01 | 3.91 | .13 |
| SAR × BBN | | | | **-7.88** | **2.57** | **.002** | -12.75 | 3.55 | < .001 |
| SAR × Post-BBN | | | | **-5.05** | **2.31** | **.029** | -7.95 | 3.21 | .014 |
| WE × Pre-BBN | | | | -4.10 | 2.81 | .15 | -4.87 | 3.89 | .21 |
| WE × BBN | | | | -0.63 | 2.56 | .81 | -5.46 | 3.53 | .12 |
| WE × Post-BBN | | | | 1.72 | 2.30 | .45 | -1.14 | 3.19 | .72 |
| Motivation × Pre-BBN | | | | 0.49 | 1.77 | .78 | 0.49 | 1.77 | .78 |
| Motivation × BBN | | | | 2.48 | 1.61 | .12 | 2.48 | 1.60 | .12 |
| Motivation × Post-BBN | | | | 0.43 | 1.45 | .77 | 0.43 | 1.45 | .77 |
| SAR × WE × Pre-BBN | | | | | | | 1.59 | 5.52 | .77 |
| SAR × WE × BBN | | | | | | | **9.93** | **5.02** | **.048** |
| SAR × WE × Post-BBN | | | | | | | 5.89 | 4.53 | .19 |

Linear mixed effects regressions for pre-ejection period. Significant main effects in Model 0 and interaction effects in Models 1 and 2 ($p < .05$) are presented in bold. [a] No-SAR = 0, SAR = 1. [b] No-WE = 0, WE = 1. [c] Reference = baseline. [d] Motivation is grand mean centered.

**Supplementary Table S12**

| Contrast | Beta | *SE* | *t*(552) | *p* |
|---|---|---|---|---|
| SAR-only vs. No-intervention (SAR x BBN effect when WE = 0) | **-12.75** | **3.55** | **-3.59** | **< .001** |
| SAR & WE vs. WE-only (SAR x BBN effect when WE = 1) | -2.83 | 3.61 | -0.78 | .43 |
| WE-only vs. No-intervention (WE x BBN effect when SAR = 0) | -5.46 | 3.53 | -1.55 | .12 |
| SAR & WE vs. SAR-only (WE x BBN effect when SAR = 1) | 4.47 | 3.62 | 1.23 | .22 |

Post-hoc analysis of the significant SAR × WE × BBN interaction for the pre-ejection period. The estimates represent the differences in the changes from baseline to BBN task of the two groups of each contrast. Significant effects ($p < .05$) are presented in bold.

### Supplementary Figure 1

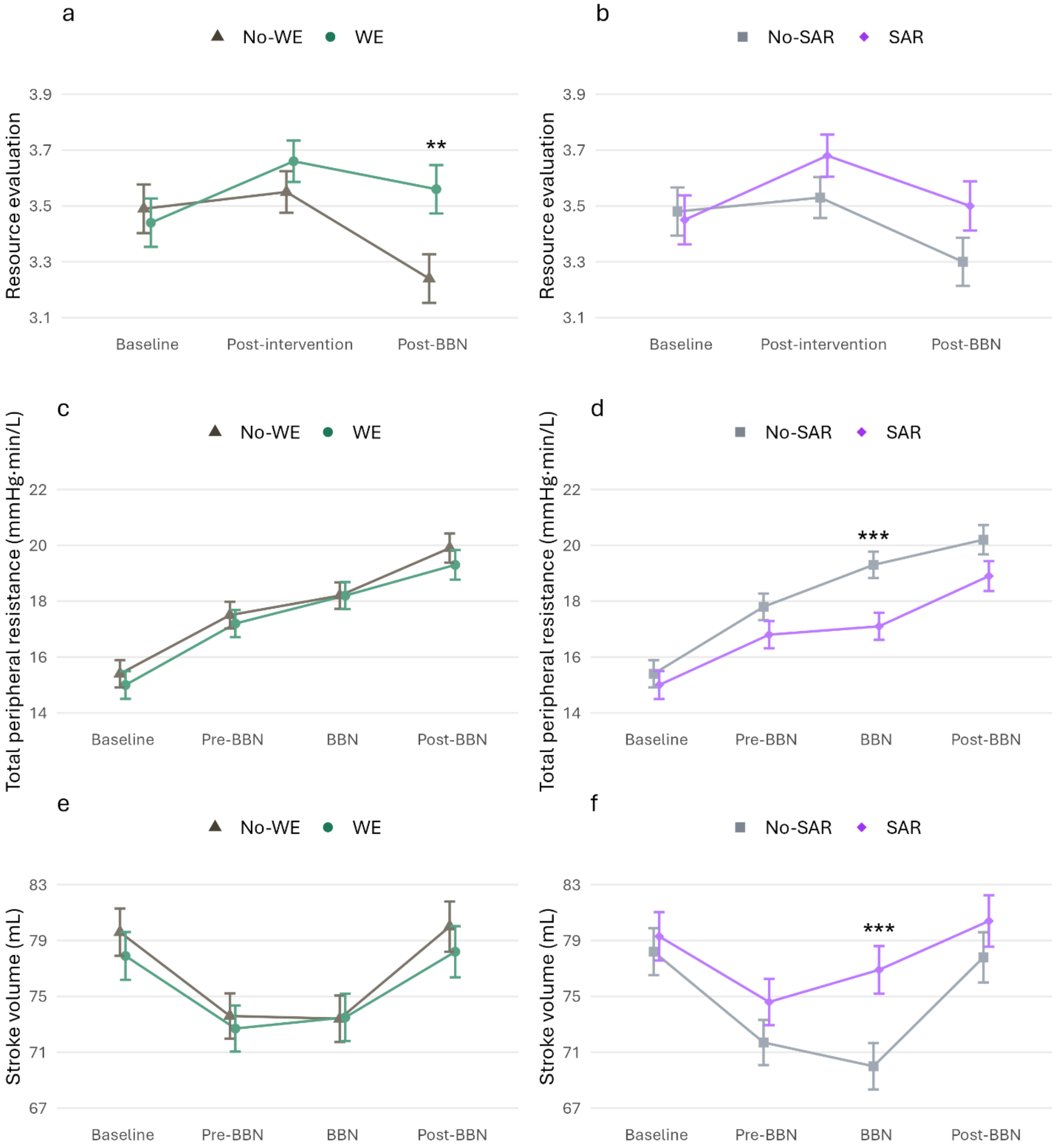


Line plots of Model 1 estimated marginal means of resource evaluation, total peripheral resistance, and stroke volume for WE vs. No-WE groups (a, c, e) and SAR vs. No-SAR groups (b, d, f). The error bar represents the standard error. Significant differences in the changes from baseline to the respective time point are marked with asterisks (** $p < .01$, *** $p < .001$).

**Supplementary Figure 2**

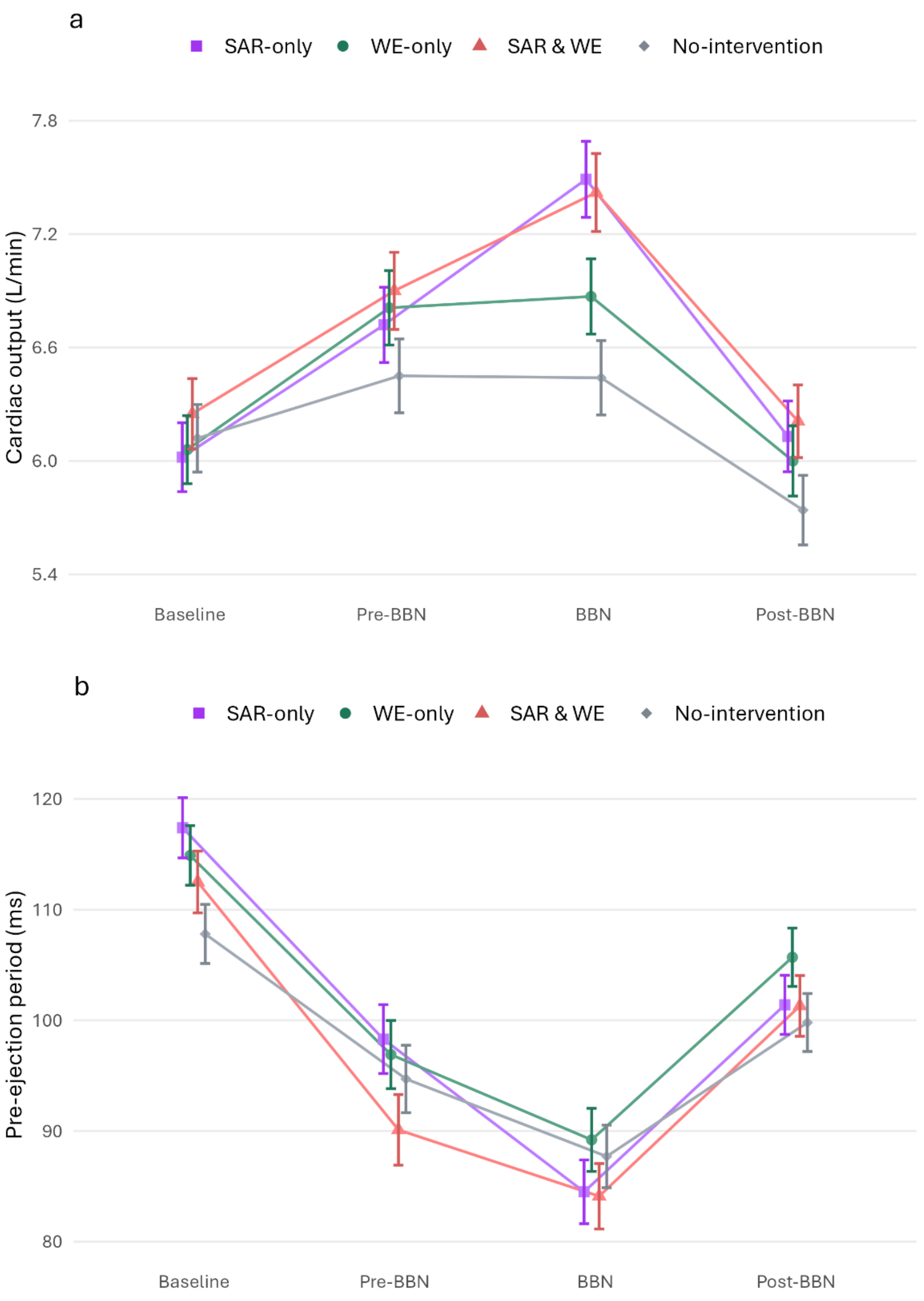


Line plots of Model 2 estimated marginal means of cardiac output (a) and pre-ejection period (b) for the SAR-only, WE-only, SAR & WE, and No-intervention groups. The error bar represents the standard error. For contrast effects, please see the Supplementary Tables S7 (CO) and S12 (PEP).
